# On Mendelian Randomisation Mixed-Scale Treatment Effect Robust Identification (MR MiSTERI) and Estimation for Causal Inference

**DOI:** 10.1101/2020.09.29.20204420

**Authors:** Zhonghua Liu, Ting Ye, Baoluo Sun, Mary Schooling, Eric Tchetgen Tchetgen

## Abstract

Standard Mendelian randomization analysis can produce biased results if the genetic variant defining instrumental variable (IV) is confounded and/or has a horizontal pleiotropic effect on the outcome of interest not mediated by the treatment. We provide novel identification conditions for the causal effect of a treatment in the presence of unmeasured confounding by leveraging an invalid IV for which both the IV independence and exclusion restriction assumptions may be violated. The proposed Mendelian Randomization Mixed-Scale Treatment Effect Robust Identification (MR MiSTERI) approach relies on (i) an assumption that the treatment effect does not vary with the invalid IV on the additive scale; and (ii) that the selection bias due to confounding does not vary with the invalid IV on the odds ratio scale; and (iii) that the residual variance for the outcome is heteroscedastic and thus varies with the invalid IV. Although assumptions (i) and (ii) have, respectively appeared in the IV literature, assumption (iii) has not; we formally establish that their conjunction can identify a causal effect even with an invalid IV subject to pleiotropy. MR MiSTERI is shown to be particularly advantageous in the presence of pervasive heterogeneity of pleiotropic effects on the additive scale. For estimation, we propose a simple and consistent three-stage estimator that can be used as preliminary estimator to a carefully constructed one-step-update estimator, which is guaranteed to be more efficient under the assumed model. In order to incorporate multiple, possibly correlated and weak IVs, a common challenge in MR studies, we develop a MAny Weak Invalid Instruments (MR MaWII MiSTERI) approach for strengthened identification and improved estimation accuracy. Both simulation studies and UK Biobank data analysis results demonstrate the robustness of the proposed MR MiSTERI method.

## 1 Introduction

Mendelian randomization (MR) is an instrumental variable (IV) approach that uses genetic variants, for example, single-nucleotide polymorphisms (SNPs), to infer the causal effect of a modifiable risk treatment on a health outcome (Smith and Ebrahim 2003). MR has recently gained popularity in epidemiological studies because, under certain conditions, it can provide unbiased estimates of causal effects even in the presence of unmeasured exposure-outcome confounding. For example, findings from a recent MR analysis assessing the causal relationship between low-density lipoprotein cholesterol and coronary heart disease (Ference et al. 2017) in an observational study are consistent with the results of earlier randomized clinical trials (Scandinavian Simvastatin Survival Study Group 1994).

For a SNP to be a valid IV, it must satisfy the following three core assumptions (Didelez and Sheehan 2007; Lawlor et al. 2008):

**(A1)** IV relevance: the SNP must be associated (not necessarily causally) with the exposure;

**(A2)** IV independence: the SNP must be independent of any unmeasured confounder of the exposure-outcome relationship;

**(A3)** Exclusion restriction: the SNP cannot have a direct effect on the outcome variable not mediated by the treatment, that is, no horizontal pleiotropic effect can be present.

The causal diagram in Figure 1(a) graphically represents the three core assumptions for a valid IV. It is well-known that even when assumptions (A1)-(A3) hold for a given IV, the average causal effect of the treatment on the outcome cannot be point identified without an additional condition, the nature of which dictates the interpretation of the identified causal effect. Specifically, Angrist et al. (1996) proved that under (A1)-(A3) and a monotonicity assumption about the IV-treatment relationship, the so-called complier average treatment effect is nonparametrically identified. More recently, Wang and Tchetgen Tchetgen (2018) established identification of population average causal effect under (A1)-(A3) and an additional assumption of no unmeasured common effect modifier of the association between the IV and the endogenous variable, and the treatment causal effect on the outcome. A special case of this assumption is that the association between the IV and the treatment variable is constant on the additive scale across values of the unmeasured confounder (Tchetgen Tchetgen et al. (2020)). In a separate strand of work, Robins (1994) identified the effect of treatment on the treated under (A1)-(A3) and a so-called “no current-treatment value interaction” assumption (A.4a) that the effect of treatment on the treated is constant on the additive scale across values of the IV. In contrast, Liu et al. (2020) established identification of the effect of treatment on the treated (ETT) under (A1)-(A3), and an assumption (A.4b) that the selection bias function defined as the odds ratio association between the potential outcome under no treatment and the treatment variable, is constant across values of the IV. Note that under the IV DAG Figure 1(a), assumption (A1) is empirically testable while (A2) and (A3) cannot be refuted empirically without a different assumption being imposed (Glymour et al. 2012). Possible violation or near violation of assumption (A1) known as the weak IV problem poses an important challenge in MR studies as the associations between a single SNP IV and complex traits can be fairly weak (Staiger and Stock 1997; Stock et al. 2002). But massive genotyped datasets have provided many such weak IVs. This has motivated a rich body of work addressing the weak IV problem under a many weak instruments framework, from a generalized method of moments perspective given individual level data (Chao and Swanson 2005; Newey and Windmeijer 2009; Davies et al. 2015), and also from a summary-data perspective (Zhao et al. 2019a,b; Ye et al. 2019). Violation of assumption (A2) can occur due to population stratification, or when a selected SNP IV is in linkage disequilibrium (LD) with another genetic variant which has a direct effect on the outcome (Didelez and Sheehan 2007). Violations of (A3) can occur when the chosen SNP IV has a non-null direct effect on the outcome not mediated by the exposure, commonly referred to as horizontal pleiotropy and is found to be widespread (Solovieff et al. 2013; Verbanck et al. 2018). A standard MR analysis (i.e. based on standard IV methods such as 2SLS) with an invalid IV that violates any of those three core assumptions might yield biased causal effect estimates.

**Figure 1:**
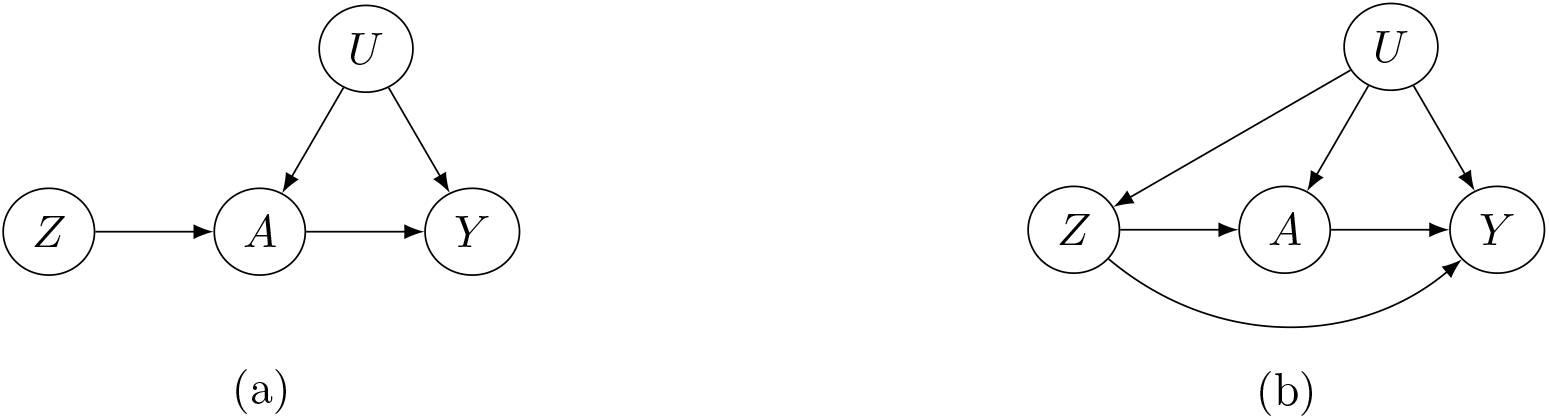
Directed acyclic graph (DAG) with an instrument (*Z*), an outcome (*Y*), a treatment (*A*) and unmeasured confounders (*U*). The left panel shows a valid Mendelian randomization study and the right panel shows violations of IV independence and exclusion restriction assumptions.

Methods to address possible violations of (A2) or (A3) given a single candidate IV are limited. Two methods have recently emerged as potentially robust against such violation under certain conditions. The first, known as MR-GxE, assumes that one has observed an environmental factor (E), which interacts with the invalid IV in its additive effects on the treatment of interest, and that such interaction is both independent of any unmeasured confounder of the exposure-outcome relationship, and does not operate on the outcome in view (Spiller et al. 2019, 2020). In other words, MR-GxE essentially assumes that while the candidate SNP (the G variable) may not be a valid IV, its additive interaction with an observed covariate constitutes a valid IV which satisfies (A1)-(A3). In contrast, MR GENIUS relies on an assumption that the residual variance of the first stage regression of the treatment on the candidate IV is heteroscedastic with respect to the candidate IV, i.e., the variance of the treatment depends on the IV, an assumption that may be viewed as strengthening of the IV relevance assumption (Lewbel 2012; Tchetgen Tchetgen et al. 2020). Interestingly, as noted by Tchetgen Tchetgen et al. (2020), existence of a GxE interaction that satisfies conditions (A1)-(A3) required by MR GxE had such an E variable (independent of *G*) been observed, necessarily implies that the heteroscedasticity condition required by MR GENIUS holds even when the relevant E variable is not directly observed. Furthermore, as it is logically possible for heteroscedasticity of the variance of the treatment to operate even in absence of any GxE interaction, MR GENIUS can be valid in certain settings where MR GxE is not.

In this paper, we develop an alternative robust MR approach for estimating a causal effect of a treatment subject to unmeasured confounding, leveraging a potentially invalid IV which fails to fulfill either IV independence or exclusion restriction assumptions. The proposed “Mendelian Randomization Mixed-Scale Treatment Effect Robust Identification” (MR MiSTERI) approach relies on (i) an assumption that the treatment effect does not vary with the invalid IV on the additive scale; and (ii) that the selection bias due to confounding does not vary with the invalid IV on the odds ratio scale; and (iii) that the residual variance for the outcome is heteroscedastic and thus varies with the invalid IV. Note that assumption (iii) is empirically testable. Although assumptions (i) and (ii) have, respectively appeared in the IV literature, assumption (iii) has not; we formally establish that their conjunction can identify a causal effect even with an invalid IV subject to pleiotropy. MR MiSTERI is shown to be particularly advantageous in the presence of pervasive heterogeneity of pleiotropic effects on the additive scale, a setting in which both MR GxE and MR GENIUS can be severely biased whenever heteroscedastic first stage residuals can fully be attributed to latent heterogeneity in SNP-treatment association (Spiller et al. 2020). For estimation, we propose a simple and consistent three-stage estimator that can be used as a preliminary estimator to a carefully constructed one-step-update estimator, which is guaranteed to be more efficient under the assumed model. In order to incorporate multiple, possibly correlated and weak IVs, a common challenge in MR studies, we develop a MAny Weak Invalid Instruments (MaWII MR MiSTERI) approach for strengthened identification and improved accuracy. Simulation study results show that our proposed MR method gives consistent estimates of the causal parameter and the selection bias parameter with nominal confidence interval coverage with an invalid IV, and the accuracy is further improved with multiple weak invalid IVs. For illustration, we apply our method to the UK Biobank data set to estimate the causal effect of body mass index (BMI) on average glucose level. We also develop an R package freely available for public use at https://github.com/zhonghualiu/MRMiSTERI.

## 2 Methods

### 2.1 Identification with a Binary Treatment and a Possibly Invalid Binary IV

Suppose that we observe data *O*_*i*_ = (*Z*_*i*_, *A*_*i*_, *Y*_*i*_) of sample size *n* drawn independently from a common population, where *Z*_*i*_, *A*_*i*_, *Y*_*i*_ denote a SNP IV, a treatment and a continuous outcome of interest for the *i*th subject (1 ≤ *i* ≤ *n*), respectively. In order to simplify the presentation, we drop the sample index *i* and do not consider observed covariates at this stage, although all the conclusions continue to hold within strata defined by observed covariates. Let *z, a, y* denote the possible values that *Z, A, Y* could take, respectively. Let *Y*_*az*_ denote the potential outcome, had possibly contrary to fact, *A* and *Z* been set to *a* and *z* respectively, and let *Y*_*a*_ denote the potential outcome had *A* been set to *a*. We are interested in estimating the ETT defined as

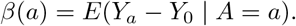

To facilitate the exposition, consider the simple setting where both the treatment and the SNP IV are binary, then the ETT simplifies to *β* = *E*(*Y*_1_ − *Y*_0_ | *A* = 1). By the consistency assumption (Hernan and Robins 2020), we know that *E*(*Y*_1_ | *A* = 1) = *E*(*Y* | *A* = 1). However, the expectation of the potential outcome *Y*_0_ among the exposed subpopulation *E*(*Y*_0_ | *A* = 1) cannot empirically be observed due to possible unmeasured confounding for the exposure-outcome relationship. The following difference captures this confounding bias on the additive scale *Bias* = *E*(*Y*_0_ | *A* = 1) − *E*(*Y*_0_ | *A* = 0), which is exactly zero only when exposed and unexposed groups are exchangeable on average (i.e. under no confounding) (Hernan and Robins 2020), and is otherwise not null. With this representation and the consistency assumption, we have

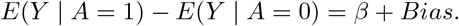

This simple equation implies that one can only estimate the sum of the causal effect *β* and the confounding bias but cannot tease them apart using the data (*A, Y*) only. With the availability of a binary candidate SNP IV *Z* that is possibly invalid, we can further stratify the population by *Z* to obtain under consistency:

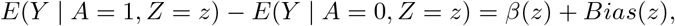

where *z* is equal to either 0 or 1, and *β*(*z*) = *E*(*Y*_*a*_ − *Y*_0_ | *A* = *a, Z* = *z*), *Bias*(*z*) = *E*(*Y*_0_ | *A* = 1, *Z* = *z*) − *E*(*Y*_0_ | *A* = 0, *Z* = *z*) denote the causal effect and bias in the stratified population with the IV taking value *z*. Note that there are only two equations but four unknown parameters: *β*(*z*), *Bias*(*z*), *z* = 0, 1. Therefore, the causal effect cannot be identified without imposing assumptions to reduce the total number of parameters to two.

Our first assumption extends the no current-treatment value interaction assumption (A.4a) originally proposed by Robins (1994), that the causal effect does not vary across the levels of the SNP IV so that *β*(*z*) is a constant as function of *z*. Formally stated:

**(B1)**Homogeneous ETT assumption: *E*(*Y*_*a*=1,*z*_ − *Y*_*a*=0,*z*_ | *A* = 1, *Z* = *z*) = *β*.

It is important to note that this assumption does not imply the exclusion restriction assumption (A3); it is perfectly compatible with presence of a direct effect of *Z* on *Y* (the direct arrow from *Z* to *Y* is present in Figure 1(b)), i.e. *E*(*Y*_*a*=0,*z*=1_ − *Y*_*a*=0,*z*=0_ |*A* = 1, *Z* = *z*) ≠ 0 as well as unmeasured confounding of the effects of Z on (A,Y), both of which we accommodate.

In order to state our second core assumption, consider the following data generating mechanism for the treatment, which encodes presence of unmeasured confounding by making dependence between *A* and *Y*_0_ explicit under a logistic model:

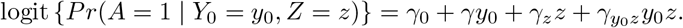

This model can of course not be estimated directly from the observed data without any additional assumption because it would require the potential outcome *Y*_0_ be observed both on the untreated (guaranteed by consistency) and the treated. Nevertheless, this model implies that the conditional (on *Z*) association between *A* and *Y*_0_ on the odds ratio scale is 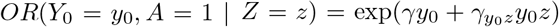. Together, *γ* and 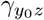 encode the selection bias due to unmeasured confounding on the log-odds ratio scale. If both *γ* and 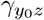 are zeros, then *A* and *Y*_0_ are conditionally (on *Z*) independent, or equivalently, there is no residual confounding bias upon conditioning on *Z*. Our second identifying assumption formally encodes assumption A4.b of Liu et al. (2020) of homogeneous odds ratio selection bias 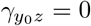:

**(B2)** Homogeneous selection bias: *OR*(*Y*_0_ = *y*_0_, *A* = *a* | *Z* = *z*) = exp(*γay*_0_).

Assumption (B2) states that the selection bias on the odds ratio scale is homogeneous across levels of the IV. Thus, this assumption allows for the presence of unmeasured confounding which, upon setting 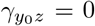 is assumed to be on average balanced with respect to the SNP IV (on the odds ratio scale). Following Liu et al. (2020), we have that under (B2)

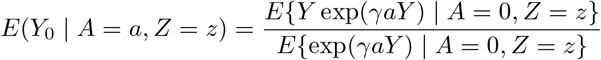

Define *ε* = *Y* − *E*(*Y* |*A, Z*), and suppose that *ε* |*A, Z* ~ *N* (0, *σ*^2^(*Z*)), then after some algebra we have,

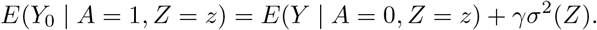

The selection bias on the odds ratio scale does not vary with the levels of the IV, however in order to achieve identification, the bias term *γσ*^2^(*Z*) must depend on the IV. This observation motivates our third assumption,

**(B3)** Outcome heteroscedasticity, that is *σ*^2^(*Z*) cannot be a constant.

Assumption (B3) is empirically testable using existing statistical methods for detecting non-constant variance in regression models. As shown by Paré et al. (2010), observed or unobserved gene–gene (GxG) and/or gene–environment (GxE) can result in changes in variance of a quantitative trait per genotype. Paré et al. (2010) found several SNPs associated with the variance of inflammation markers. Therefore, GxE interaction can be inferred from genetic variants associated with phenotypic variability without the need of measuring environmental factors, usually termed variance quatitative trait loci (vQTL) screening (Marderstein et al. 2021). In particular, Wang et al. (2019) performed a genome-wide vQTL analysis of about 5.6 million variants on 348,501 unrelated individuals of European ancestry in the UK Biobank and identified 75 significant vQTLs for 9 quantitative traits (out of 13 traits under study). Hence, one can generally expect this novel identification assumption (B3) to hold in the presence of pervasive heterogeneity of pleiotropic effects on the additive scale for the outcome, a setting in which both MR GxE and MR GENIUS can be severely biased. Under assumptions (B1)-(B3), we have

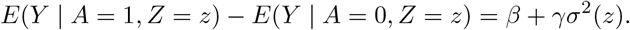

Note that *σ*^2^(*z*) is the variance of Y given z, and thus can be estimated with no bias. For the binary IV *Z, σ*^2^(*Z* = 0) and *σ*^2^(*Z* = 1) are the variance of *Y* within each IV stratum and can easily be estimated using the sample variance in each group. We will describe estimating procedures in the next section for more general settings where *Z* is not necessarily binary. Importantly, in the present case, we now have two equations and two unknown parameters *β, γ*. Denote *D*(*Z* = *z*) = *E*(*Y* | *A* = 1, *Z* = *z*) − *E*(*Y* | *A* = 0, *Z* = *z*). We have the following main identification result.

#### Theorem 1.

*Under assumptions (B1)-(B3), the selection bias parameter and the causal effect of interest are uniquely identified as follows:*

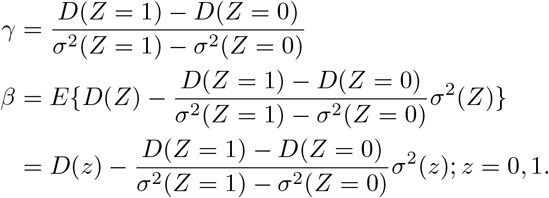

In an observed sample, one can simply use the sample versions of unknown quantities to obtain estimates for *β* and *γ* respectively. Standard errors can be deduced by a direct application of the multivariate delta method, or by using resampling techniques such as the jackknife or the bootstrap. **Remark 1**. Note that as stated in the theorem, the causal parameter *β* can be estimated based on data among either *Z* = 0 or *Z* = 1 group. In practical settings, in order to improve efficiency, one may take either unweighted average of the two estimates as given in the theorem, or their inverse variance weighted average. In fact, one may fit a saturated linear model for the variance *σ*^2^(*Z* = *z*) = *b*_0_ + *bz*; We give a graphical illustration of our identification strategy as shown in Figure 2. By using the outcome heteroscedasticity assumption (B3), we can identify the selection bias parameter *γ* and then the causal effect parameter *β*. Without the assumption (B3) as shown by the red dashed line, we can only obtain *β* + *γb*_0_ and thus cannot tease apart the causal effect and the selection bias.

**Figure 2:**
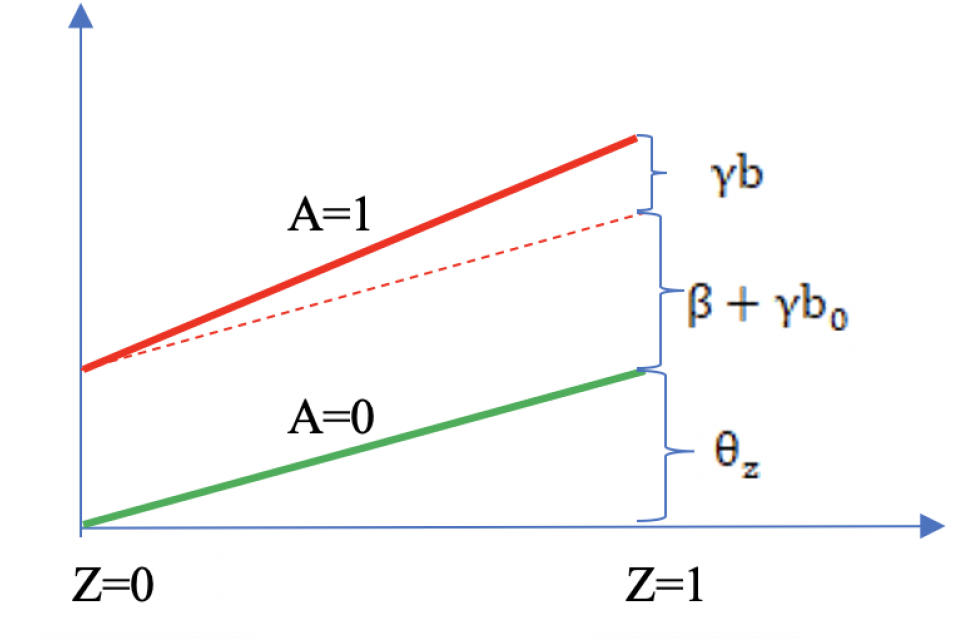
An illustration of our identification strategy for a binary treatment *A* and binary IV *Z*. The green line represents *E*(*Y* |*A* = 0, *Z* = *z*) and its difference between *Z* = 1 and *Z* = 0 is *θ*_*z*_. The red dashed line (parallel to the green line) represents the hypothetical setting when *σ*^2^(*Z* = *z*) is constant, and the red solid line represents the setting when *σ*^2^(*Z* = *z*) is not a constant function in *Z*. Note that *b*_0_ and *b* are treated as known.

### 2.2 Identification with a Continuous Treatment and a Possibly Invalid Discrete IV

The identification strategy in the previous section extends to the more general setting of a continuous treatment and a more general discrete IV, for example, the SNP IV taking values 0, 1, 2 corresponding to the number of minor alleles. To do so, we introduce the notion of generalized conditional odds ratio function as a measure of the conditional association between a continuous treatment and a continuous outcome. To ground idea, consider the following model for the treatment free potential outcome:

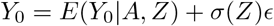

where *ϵ* is independent standard normal; then one can show that the generalized conditional odds ratio function associated with *Y*_0_ and *A* given *Z* with (*Y*_0_ = 0, *A* = 0) taken as reference values (Chen 2007) is given by

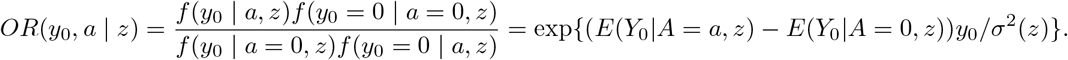

By Assumption (B2), we have

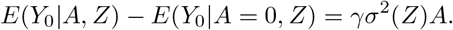

Therefore,

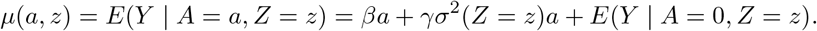

For practical interpretation, we assume *A* has been centered so that *A* = 0 actually represents average treatment value in the study population. Assume the following model *E*(*Y* | *A* = 0, *Z* = *z*) = *θ*_0_ + *θ*_*z*_*z* and a log linear model for *σ*^2^(*Z*) such that

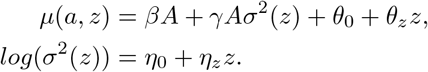

The conditional distribution of *Y* given *A* and *Z* is

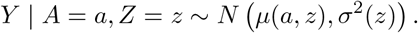

Then we can simply use standard maximum likelihood estimation (MLE) to obtain consistent and fully efficient estimates for all parameters under the assumed model. However, the likelihood may not be log-concave and direct maximization of the likelihood function with respect to all parameters might not always converge. Instead, we propose a novel three-stage estimation procedure which can provide a consistent but inefficient preliminary estimator of unknown parameters. We then propose a carefully constructed one-step update estimator to obtain a consistent and fully efficient estimator. Our three-stage estimation method works as follows.

**Stage 1:** Fit the following linear regression using standard (weighted) least-squares

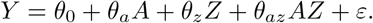

We obtain parameter estimates 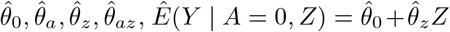, with corresponding estimated residuals 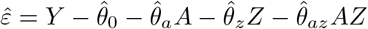.

**Stage 2:** Regress the squared residuals 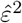 on (1, *Z*) in a log-linear model and obtain parameter estimates 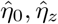 and 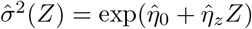.

**Stage 3:** Regress *Y* − *Ê*(*Y* | *A* = 0, *Z*) on *A* and 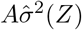 without an intercept term and obtain the two corresponding regression coefficients estimates 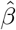 and 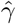 respectively. Alternatively, we may also fit 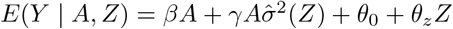.

The proposed three-stage estimation procedure is computationally convenient, appears to always converge and provides a consistent estimator for all parameters under the assumed model. Next we propose the following one-step update to obtain a fully efficient estimator. Consider the following normal model

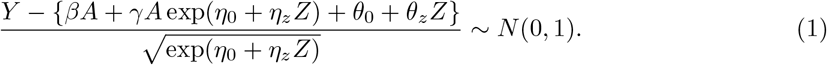

Denote Θ = (*β, γ, η*_0_, *η*_*z*_, *θ*_0_, *θ*_*z*_) and denote the log-likelihood function as *l*(Θ; *Y, A, Z*) for an individual. With a sample of size *n*, the log-likelihood function is

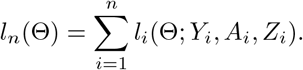

The score function *S*_*n*_(Θ) = *∂l*_*n*_(Θ)*/∂*Θ is defined as the first order derivative of the log-likelihood function with respect to Θ, and use *i*_*n*_(Θ) = *∂*^2^*l*_*n*_(Θ)*/∂*Θ*∂*Θ^*T*^ to denote the second order derivative of the log-likelihood function. Suppose we have obtained a consistent estimator 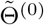 of Θ using the three-stage estimating procedure, then one can show that a consistent and fully efficient estimator is given by the one-step update estimator 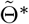

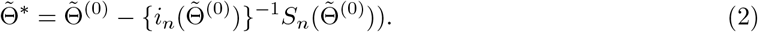

**Remark 2**. In biomedical studies, continuous outcomes of interest are often approximately normally distributed, and the normal linear regression is also routinely used by applied researchers.

When the normal assumption is violated, a typical remedy is to apply a transformation to the outcome prior to modeling it, such as the log-transformation or the more general Box-Cox transformation to achieve approximate normality. Alternatively, in section 2.4 we propose to model the error distribution as a more flexible Gaussian mixture model which is more robust than the Gaussian model. Alternatively, as described in the Supplemental Materials, one may consider a semiparametric location-scale model which allows the distribution of standardized residuals to remain unrestricted, a potential more robust approach although computationally more demanding.

### 2.3 Estimation and Inference under Many Weak Invalid IVs

Weak identification bias is a salient issue that needs special attention when using genetic data to strengthen causal inference, as most genetic variants typically have weak association signals. When many genetic variants are available, we recommend using the conditional maximum likelihood estimator (CMLE) based on (1), where *Z* is replaced with a multi-dimensional vector. Let 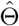 be the solution to the corresponding score functions, i.e., 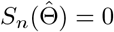; let

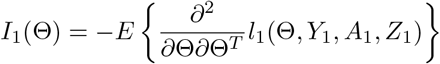

be the Fisher information matrix based on the conditional likelihood function for one observation. Let *k* be the total number of parameters in Θ, which is equal to 4 + 2*p* when *Z* is *p* dimensional. When *λ*_min_ {*nI*_1_(Θ)}*/k* → ∞ with *λ*_min_ {*nI*_1_(Θ)} being the minimum eigenvalue of *nI*_1_(Θ), we show in the Supplementary Materials (Theorem 2) that 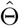 is consistent and asymptotically normal as the sample size goes to infinity, and satisfies

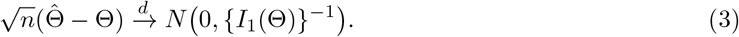

In other words, the CMLE is robust to weak identification bias and the usual inference procedure can be directly applied.

The key condition *λ*_min_{*nI*_1_(Θ)}*/k* → ∞ warrants more discussion. Note that the quantity *λ*_min_ {*nI*_1_(Θ)}*/k* measures the ratio between the amount of information that a sample of size *n* carries about the unknown parameter and the number of parameters. In classical strong identification scenarios where the minimum eigenvalue of *I*_1_(Θ) is assumed to be lower bounded by a constant, then the condition simplifies to that the number of parameter is small compared with sample size, i.e., *n/k* → ∞. However, when identification is weak, the minimum eigenvalue of *I*_1_(Θ) can be small. In practice, the condition *λ*_min_{*nI*_1_(Θ)}*/k* → ∞ can be evaluated by

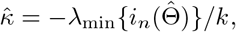

which is the ratio between the minimum eigenvalue of the negative Hessian matrix and the number of parameters. We remark that the condition stated in the Supplementary Materials (Theorem 2) is more general and applies to general likelihood-based methods, which, for example, implies the condition for consistency for the profile likelihood estimator (MR.raps) in Zhao et al. (2019b). In practice, we recommend checking that the 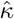 is at least greater than 10 based on our simulation studies. For valid statistical inference, a consistent estimator for the variance covariance matrix for 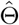 is simply the negative Hessian matrix 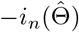, whose corresponding diagonal element estimates the variance of the treatment effect estimator 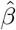. Other variants of the CMLE can also be used, for example, the one-step iteration estimator 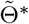 in (2) is asymptotically equivalent to the CMLE 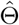 as long as the initial estimator 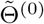 is 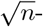 consistent (Shao 2003).

### 2.4 More Robust Gaussian Mixture Model for the Outcome

As mentioned in Remark 2, although many continuous outcomes can be well modelled using normal linear model, however sometimes, a more robust model for the outcome error term is desired. As shown by Verbeke and Lesaffre (1996), a Gaussian mixture model is more general and thus more robust than the normal model for modelling an outcome error distribution. Consider the general location-scale model

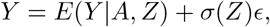

where *ϵ* ⊥ *A, Z*. Under assumptions (B1)-(B3), the conditional mean function is given by

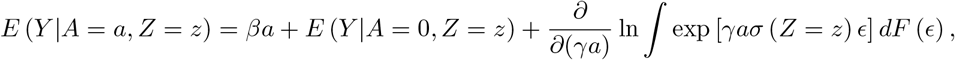

where *F* (·) denotes the cumulative distribution function of *ϵ*. We assume that the outcome error distribution can be reasonably approximated by a Gaussian mixture model with enough components (Verbeke and Lesaffre 1996). Specifically, let 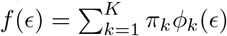 where *ϕ*_*k*_(·) is the normal density with mean *µ*_*k*_ and variance 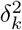 satisfying the constraints

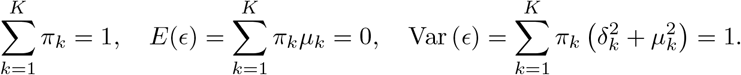

The conditional mean function with Gaussian mixture error is given by

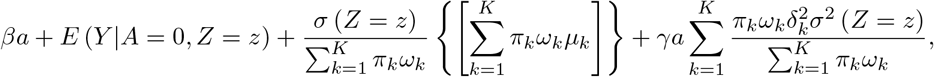

Where 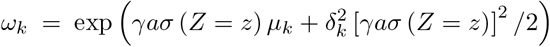. The conditional distribution is 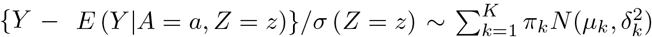. In practice, estimation of (*β, γ*), which are of primary interest, may proceed under a user-specified integer value *K <* as well as parametric models for *E*(*Y* |*A* = 0, *Z* = *z*) and *σ*(*Z* = *z*) via an alternating optimization algorithm which we describe in the Supplementary Materials.

## 3 Simulation Studies

In this section, we conduct extensive simulation studies to evaluate the finite-sample performance of MR-MiSTERI compared to its main competitors, standard two stage least squares (TSLS) and MR GENIUS. We considered sample sizes of *n* = 10000, 30000, 100000. The IV *Z* is generated from a Binomial distribution with probability equal to 0.3 (minor allele frequency). The treatment *A* is generated from a standard normal distribution *N* (0, 1). The outcome *Y* is also generated from a normal distribution with mean and variance compatible with assumptions B1-B3:

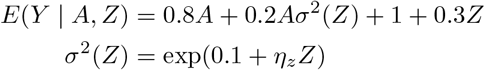

where *η*_*z*_ is set to be 0.2, 0.15, 0.1, 0.05. Under this model, we have that the ETT is equal to *β* = 0.8 and *γ* = 0.2. We generated in total 1000 such data sets and applied our proposed method to each of them along with TSLS and MR GENIUS. We found that TSLS gives severely biased estimates of the causal effect with estimates as extreme as −195.72 and standard errors as large as 381.57. MR GENIUS is also severely biased as the average point estimate for the causal effect is −14.99. Numerical results for TSLS and MR GENIUS are expected as their corresponding identification conditions are not met in the simulation setup. Specifically, both exclusion restriction and heteroscedasticity of the treatment with respect to *Z* assumptions do not hold. Simulation results for MR MiSTERI are summarized in Table 1. We found that when sample size is 10000, the bias and standard error of the causal effect and selection bias parameter estimates become larger when the IV strength *η*_*z*_ decreases from 0.2 to 0.1. The causal effect is less sensitive to the IV strength compared to the selection bias parameter *γ*. As we increase sample size, the bias decreases and becomes negligible at n=100000. Confidence intervals achieved the nominal 95% coverage.

**Table 1:**
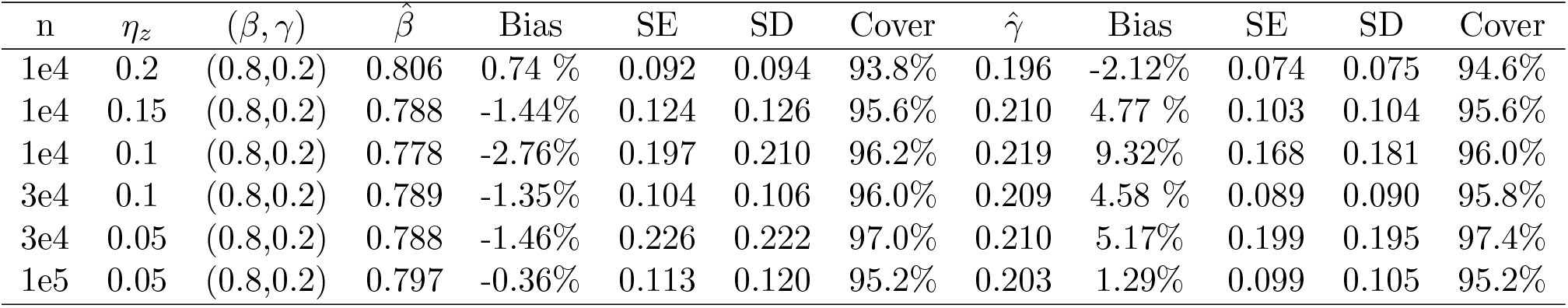
Simulation results for estimating the confounding bias parameter *γ* and the ETT *β* using the one-step estimator approach (without covariates) based on 1000 Monte Carlo experiments with a range of sample sizes n. 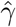 is the averaged point estimates of *γ*, and bias is calculated in percentage format. Likewise for 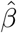. SE is the averaged standard error. SD stands for the sample standard deviation of the 1000 point estimates for *γ* and *β*. Coverage is calculated as the proportion of 95% confidence intervals that contains the true parameter value among those 1000 experiments. We vary the value of *η*_*z*_ in *σ*^2^(*Z*) = *exp*(*η*_0_ + *η*_*z*_*Z*)) to assess the impact of decreasing IV strength on the estimation results.

The second simulation study is designed to evaluate the finite sample performance of the proposed three-stage estimator and the CMLE in the presence of many weak invalid IVs. The sample size is set to be 100000, each IV *Z*_*j*_, *j* = 1, …, *p*, is still generated to take values 0,1,2 with minor allele frequency equal to 0.3. The treatment *A* is generated from standard normal distribution.

The outcome *Y* is generated from a normal distribution with mean and variance under B1-B3:

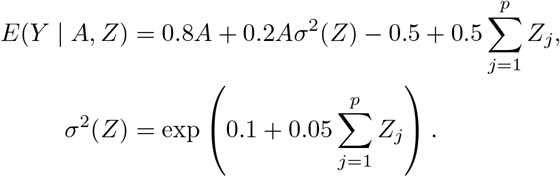

Simulation results are presented in Table 2, where the standard error (SE) for the three-stage estimator is the bootstrap estimator approximated using 100 Monte Carlo simulations, the SE for the CMLE is obtained from the inverse Hessian matrix discussed in Section 2.3. From results in Table 2, the three-stage estimator has negligible bias when *p* = 20, but shows evidently larger bias when *p* grows to 50, which is also reflected by the fact that coverage of 95% CI is substantially lower than its nominal level. In contrast, the CMLE shows negligible bias in both scenarios. The standard errors for the CMLE are close to Monte Carlo standard deviation, and the CMLE estimators show nominal coverage in both simulation scenarios. These observations agree with our theoretical assessment that the CMLE is efficient and is robust to many weak invalid IVs. In particular, it is consistent and asymptotically normal as long as the condition 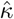 is reasonably large. Based on empirical evaluations (in the Supplementary Materials), we recommend checking that the 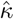 value is at least greater than 10.

**Table 2:**
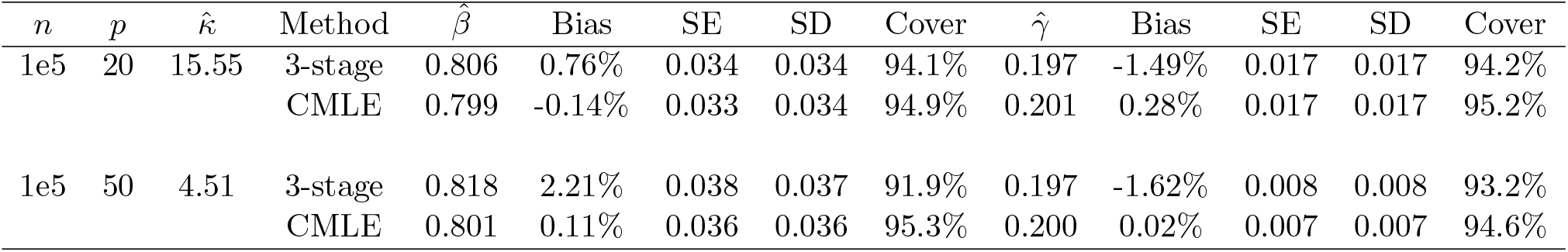
Simulation results for estimating the confounding bias parameter *γ* = 0.2 and the ETT *β* = 0.8 using the three-stage estimator and the CMLE based on 1000 Monte Carlo experiments, with *n* = 100000 and varying *p*. The third column is the averaged 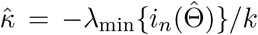, which is the consistency and asymptotic normality condition for the CMLE and is preferably to be large. 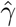 is the averaged point estimates of *γ*, and bias is calculated in percentage format. Likewise for 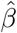. SE is the averaged standard error. SD stands for the sample standard deviation of the 1000 point estimates for *γ* and *β*. Coverage is calculated as the proportion of 95% confidence intervals that contains the true parameter value among those 1000 experiments.

We similarly generate the treatment *A* from the standard normal distribution, the IV *Z* which takes values in {0, 1, 2} with minor allele frequency equal to 0.3. However we generate *Y* = *E*(*Y* | *A,Z*) + *σ*(*Z*)*ϵ* where 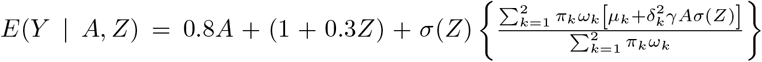, *σ*^2^(*Z*) = exp(0.1 + *η*_*z*_*Z*), and the error *ϵ* is generated from a Gaussian mixture distribution with two components,

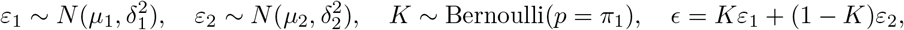

with the parameter values (*π*_1_, *π*_2_) = (0.4, 0.6), (*µ*_1_, *µ*_2_) = (−0.6, 0.4) and (*δ*_1_, *δ*_2_) = (0.5, 1.049). We vary the value of *η*_*z*_ in the set 0.1, 0.25, 0.5 to assess the impact of IV strength. The results using the proposed Gaussian mixture method with *K* = 2 are summarized in Table 3. There is noticeable finite-sample bias and under-coverage when *η*_*z*_ = 0.1, especially for estimation of the selection bias parameter *γ*, but the performance improves with larger *η*_*z*_ or sample size.

**Table 3:** Simulation results for estimating the confounding bias parameter *γ* = 0.2 and the ETT *β* = 0.8 under Gaussian mixture error based on 1000 replicates. See the caption of Table 1 for description of the summary statistics.

| $n$ | $\eta_z$ | $\hat{\beta}$ | Bias | SE | SD | Cover | $\hat{\gamma}$ | Bias | SE | SD | Cover |
| --- | --- | --- | --- | --- | --- | --- | --- | --- | --- | --- | --- |
| 1e4 | 0.10 | 0.771 | -3.64% | 0.373 | 0.195 | 75.6% | 0.256 | 27.78% | 0.314 | 0.211 | 66.7% |
|  | 0.25 | 0.798 | -0.29% | 0.056 | 0.059 | 93.9% | 0.205 | 2.39% | 0.044 | 0.049 | 91.4% |
|  | 0.50 | 0.799 | -0.12% | 0.039 | 0.039 | 95.9% | 0.201 | 0.69% | 0.026 | 0.026 | 96.0% |
| 3e4 | 0.10 | 0.780 | -2.54% | 0.106 | 0.084 | 77.0% | 0.230 | 14.95% | 0.090 | 0.101 | 69.2% |
|  | 0.25 | 0.799 | -0.09% | 0.032 | 0.033 | 95.1% | 0.202 | 0.80% | 0.025 | 0.027 | 92.9% |
|  | 0.50 | 0.799 | -0.07% | 0.023 | 0.022 | 95.4% | 0.201 | 0.36% | 0.015 | 0.015 | 95.8% |
| 1e5 | 0.10 | 0.793 | -0.86% | 0.031 | 0.041 | 78.0% | 0.210 | 4.96% | 0.026 | 0.052 | 69.5% |
|  | 0.25 | 0.800 | 0.03% | 0.018 | 0.018 | 94.7% | 0.200 | 0.24% | 0.014 | 0.015 | 93.5% |
|  | 0.50 | 0.800 | 0.03% | 0.012 | 0.012 | 95.3% | 0.200 | 0.14% | 0.008 | 0.008 | 95.6% |

## 4 An Application to the Large-Scale UK Biobank Study Data

UK Biobank is a large-scale ongoing prospective cohort study that recruited around 500,000 participants aged 40-69 in 2006-2010. Participants provided biological samples, completed questionnaires, underwent assessments, and had nurse led interviews. Follow up is chiefly through cohort-wide linkages to National Health Service data, including electronic, coded death certificate, hospital and primary care data (Sudlow et al. 2015). To control for population stratification, we restricted our analysis to participants with self-reported and genetically validated white British ancestry. For quality control, we also excluded participants with (1) excess relatedness (more than 10 putative third-degree relatives) or (2) mismatched information on sex between genotyping and self-report, or (3) sex-chromosomes not XX or XY, or (4) poor-quality genotyping based on heterozygosity and missing rates *>* 2%.

To illustrate our methods, we extracted a total of 289 010 white British subjects from the UK Biobank data with complete measurements in body mass index (BMI) and glucose levels. We further selected BMI associated SNP potential IVs among the list provided by Sun et al. (2019). We found seven SNPs (in Table 4) that are predictive of the residual variance of glucose levels in our Stage 2 regression (with p-values *P <* 0.01), that is, those seven SNPs might satisfy assumption (B3). The average BMI was 27.39 *kg/m*^2^ (SD: 4.75 *kg/m*^2^), and the average glucose level was 5.12 *mmol/L* (SD: 1.21 *mmol/L*). We applied MR MiSTERI to this data with the goal of evaluating the causal relationship between BMI as treatment and glucose level as outcome; analysis results are summarized in Table 4.

**Table 4:** UK Biobank data analysis results. Columns 2-5 contain the results based on the model (1), columns 6-9 contain results based on the Gaussian mixture model described in Section 2.4, columns 10-11 contain results using the standard two-stage least squares using the R package *ivreg*.

| SNP | $\hat{\beta}$ | $se(\hat{\beta})$ | $\hat{\gamma}$ | $se(\hat{\gamma})$ | $\hat{\beta}^*$ | $se(\hat{\beta}^*)$ | $\hat{\gamma}^*$ | $se(\hat{\gamma}^*)$ | $\hat{\beta}^{**}$ | $se(\hat{\beta}^{**})$ |
| --- | --- | --- | --- | --- | --- | --- | --- | --- | --- | --- |
| rs3736485 | 0.041 | 0.013 | 0.0001 | 0.0094 | 0.027 | 0.001 | 0.0031 | 0.0002 | 0.200 | 0.068 |
| rs1558902 | 0.023 | 0.006 | 0.0128 | 0.0046 | 0.040 | 0.001 | 0.0025 | 0.0002 | 0.064 | 0.009 |
| rs1808579 | 0.041 | 0.014 | 0.0001 | 0.0098 | 0.026 | 0.001 | 0.0042 | 0.0002 | 0.107 | 0.038 |
| rs13021737 | 0.051 | 0.012 | -0.0065 | 0.0084 | 0.036 | 0.001 | -0.0043 | 0.0005 | 0.030 | 0.016 |
| rs2176040 | 0.041 | 0.014 | 0.0585 | 0.0099 | 0.026 | 0.001 | 0.0009 | 0.0003 | -1.619 | 2.041 |
| rs10968576 | 0.047 | 0.016 | -0.0040 | 0.0111 | 0.046 | 0.001 | -0.0005 | 0.0004 | 0.079 | 0.025 |
| rs10132280 | 0.030 | 0.013 | 0.0081 | 0.0094 | 0.062 | 0.004 | -0.0228 | 0.0005 | 0.049 | 0.029 |
| Allele score | 0.030 | 0.003 | 0.0078 | 0.0021 | 0.033 | 0.005 | 0.0105 | 0.0002 | 0.089 | 0.009 |

For comparison purposes, we also include analysis results for standard two-stage least square (TSLS) implemented in the R package *ivreg* (Fox et al. 2020). The allele score for the seven SNPs selected is defined as the signed sum of their minor alleles, where the sign is determined by the regression coefficient in our stage 2 regression. We also implemented MR-GENIUS method (Tchetgen Tchetgen et al. 2020) using the same seven SNPs, the causal effect of BMI on glucose was estimated as 0.046 (se: 0.01, 95% CI: (0.0264,0.0656), *P*-value: 2.24 × 10^−6^). Crude regression analysis by simply regressing glucose on BMI gives an estimate of 0.041 (se: 4.65 × 10^−4^, 95% CI: (0.0401, 0.0419), *P*-value: *<* 2 × 10^−16^). Further, we included all seven SNPs into a conventional regression together with BMI, and obtained a similar estimate 0.039 (se: 4.65 × 10^−4^, 95% CI: (0.0381, 0.0399), *P*-value: *<* 2 × 10^−16^). It is interesting to consider results obtained by selecting each SNP as single candidate IV as summarized in the table. For instance, take the SNP rs2176040, the corresponding causal effect estimate is 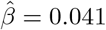 (se: 0.0139, 95% CI: (0.0138, 0.0682), *P*-value: 0.003), and the selection bias parameter is estimated as 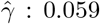 (se: 0.0098, 95% CI: (0.0398, 0.0782), *P*-value: 2.93 × 10^−9^). However, the TSLS method gives causal effect estimate −1.619 (se: 2.04, 95% CI: (−5.6174, 2.3794), *P*-value: 0.428). These conflicting findings may be due to SNP rs2176040 being an invalid IV, in which case TSLS likely yields biased results. We further combine 20 SNPs, 13 of which are weakly associated with the residual variance of glucose levels, we obtain using CMLE a causal effect estimate of BMI on glucose 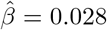 (se: 0.0025, 95% CI: (0.0231, 0.0329), *P*-value: 6.87 × 10^−31^), and a selection bias estimate of 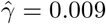 (se: 0.0017, 95% CI: (0.0057, 0.0123), *P*-value: 2.82 × 10^−7^). The consistency and asymptotic normality condition 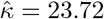, greater than our recommended value 10. This final analysis suggests a somewhat smaller treatment effect size than all the other MR methods, while providing strong statistical evidence of a positive effect with a relatively small but statistically significant amount of confounding bias detected.

## 5 Discussion

In this paper, we have proposed a novel MR MiSTERI method for identification and inference about causal effects in observational studies. MR MiSTERI leverages a possible association between candidate IV SNPs and the variance of the outcome in order to disentangle confounding bias from the causal effect of interest. Importantly, MR MiSTERI can recover unbiased inferences about the causal effect in view even when none of the candidate IV SNPs is a valid IV. Key assumptions which the approach relies on include no additive interaction involving a candidate IV SNP and the treatment causal effect in the outcome model, and a requirement that the amount of selection bias (measured on the odds ratio scale) remains constant as a function of SNP values. Violation of either assumption may result in incorrect inferences about treatment effect. Although not pursued in this paper, robustness to such violation may be assessed via a sensitivity analysis in which the impact of various departures from the assumption may be explored by varying sensitivity parameters. Alternatively, as illustrated in our application, providing inferences using a variety of MR methods each of which relying on a different set of assumptions provides a robustness check of empirical findings relative to underlying assumptions needed for a causal interpretation of results. We did not include the MR GxE method for numerical comparison because it requires users to specify an observed covariate interacting with the SNP IV. For a particular data analysis problem as we did in Section **??**, we typically do not have sufficient prior knowledge about which covariate (out of thousands) might interact with a given SNP. In contrast, MR GENIUS method requires that the treatment variable admits a residual which is heteroscedastic with respect to the IV, while MR MiSTERI requires that the outcome admits a residual that is heteroscedastic with respect to the IV. In practice, one can in principle perform a formal statistical test to determine which method might be most suitable for a particular data analysis problem. Note that both MR MiSTERI and MR GENIUS methods do not require us to specify any covariate a priori interacting with the candidate SNP IV, and hence are more convenient for routine data analyses.

A notable robustness property of CMLE estimator of MR MiSTERI is that as we formally establish, it is robust to many weak IV bias, thus providing certain theoretical guarantees of reliable performance in many common MR settings where one might have available a relatively large number of weak candidate IVs, many of which may be subject to pleiotropy. While the proposed methods require a correctly specified Gaussian model for the conditional distribution of *Y* given *A* and *Z*, such an assumption can be relaxed. In fact, we also consider a Gaussian mixture model as a framework to assess and possibly correct for potential violation of this assumption. Furthermore, in the Supplementary Materials, we briefly describe a semiparametric three-stage estimation approach under the location-scale model which allows the distribution of standardized residuals to remain unrestricted. It is of interest to investigate robustness and efficiency properties of this more flexible estimator, which we plan to pursue in future work. It is likewise of interest to investigate whether the proposed methods can be extended to the important setting of a binary outcome, which is of common occurrence in epidemiology and other health and social sciences.

## Data Availability

This research has been conducted using the UK Biobank resource (https://www.ukbiobank.ac.uk) under application number 44430.

## Acknowledgements

Dr. Zhonghua Liu’s research is supported by the Start-up research fund (000250348) of the University of Hong Kong. Dr. Eric Tchetgen Tchetgen is supported by grants R01AG065276, R01CA222147 and R01AI27271. Dr. Baoluo Sun is supported by the National University of Singapore Start-up Grant (R-155-000-203-133). This research has been conducted using the UK Biobank resource (https://www.ukbiobank.ac.uk) under application number 44430. Conflict of interest: None declared.

## Supplementary Materials

### Proof of Theorem 1

*Proof*. Under assumptions (B1) - (B3), we have

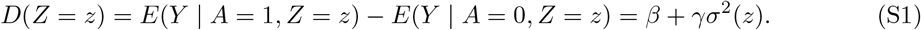

Hence, we have the following two equations

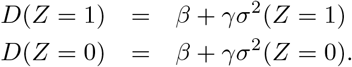

Solving these two equations simultaneously gives the result in Theorem 1.□

## Estimation and Inference under Weak Identification

Consider the following normal model also given in the main text

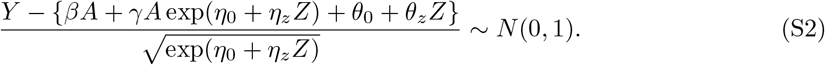

Let

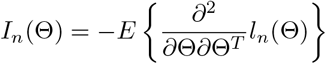

be the Fisher information matrix for a sample of size *n, λ*_min_{*I*_*n*_(Θ)} be the minimum eigenvalue of *I*_*n*_(Θ), *tr*(*A*) be the trace of a square matrix *A*.

Condition 2 summaries the regularity conditions.

**Condition 2** (Regularity Conditions). *For every o* = (*y, a, z*) *in the support of O* = (*Y, A, Z*), *the likelihood function denoted by f* (Θ *o*) *is twice continuously differentiable in* Θ *and satisfies*

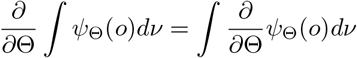

*for ψ*_Θ_(*o*) = *f*_Θ_(*o*) *and* = *∂f*_Θ_(*o*)*/∂*Θ, *where ν is a suitable measure; there is a positive ϵ such that the Fisher information matrix I*_*n*_(Γ) *is positive definite for* Γ : ‖ Γ− Θ ‖ *< ϵ and for any given* Θ, *there exists a positive number ϵ and a positive integrable function h, such that*

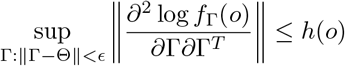

*for all o in the support of O, where* 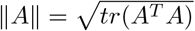 *for any matrix A*.

Theorem 3 establishes the consistency and asymptotic normality of solutions to the score function.

### Theorem 3.

*(a) Under Condition 2 and assume that the observed data O*_*i*_, *i* = 1, …, *n are in-dependent and identically distributed, if k/*[*λ*_min_{*nI*_1_(Θ)}] → 0, *then the sequence of conditional maximum likelihood estimators* 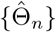 *is consistent, i*.*e*.,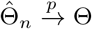.

*(b) Any consistent sequence* 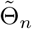*such that* 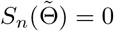 *is asymptotically normal, and as n* → ∞,

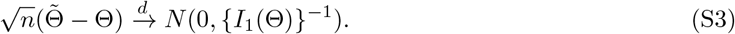

In fact, the general consistency condition in Theorem 3 can be written as *λ*_min_{*nI*_1_(Θ)} → ∞ and

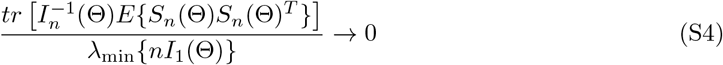

for general likelihood-based methods, which, for example, applies to the profile-likelihood estimator in Zhao et al. (2019b). Specifically, in Zhao et al. (2019b), in their notations, *k* = 1, *E* {*S*_*n*_(Θ)*S*_*n*_(Θ)^*T*^} = *V*_1_, *I*_*n*_(Θ) = *V*_2_, and 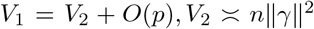, and thus, condition (S4) becomes 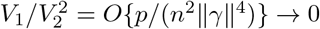, which is the condition for consistency in Zhao et al. (2019b, Theorem 3.1).

*Proof*. First, notice that the condition *k/*[*λ*_min_{*nI*_1_(Θ)}] → 0 implies *λ*_min_{*nI*_1_(Θ)} → ∞. Let *B*_*n*_(*c*) = {Γ : ‖ [*I*_*n*_(Θ)]^1*/*2^(Γ − Θ) ‖ ≤ *c*} for *c >* 0, where 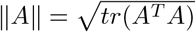 for any matrix *A*. Since *B*_*n*_(*c*) shrinks to Θ as *n* → ∞, the consistency result in Theorem 3(a) is implied by the fact that for any *e >* 0, there exists *c >* 0 and *n*_0_ *>* 1 such that

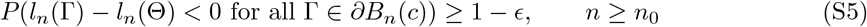

and 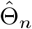 is unique for *n* ≥*n*_0_, where *∂B*_*n*_(*c*) is the boundary of *B*_*n*_(*c*). For Γ ∈*∂B*_*n*_(*c*), the Taylor expansion gives

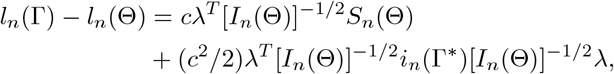

where *λ* = [*I*_*n*_(Θ)]^1*/*2^(Γ ™ Θ)*/c* satisfying ‖*λ‖* = 1, *i*_*n*_(Γ) = *∂*^2^*l*_*n*_(Γ)*/∂*Γ*∂*Γ^*T*^, and Γ^∗^ lies between Γ and Θ. Note that

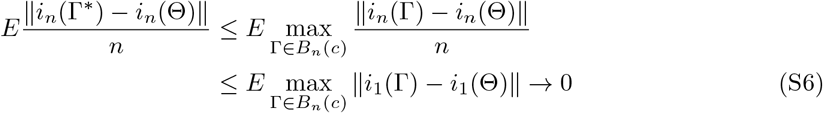

which follows from the dominated convergence theorem combined with the facts that (a) *i*_1_(Γ) = *∂l*_1_(Γ)*/∂*Γ*∂*Γ^*T*^ is continuous in a neighborhood of Θ for any fixed observation; (b) *B*_*n*_(*c*) shrinks to {Θ} from *λ*_min_(*I*_*n*_(Θ)) → ∞; and (c) for sufficiently large *n*, 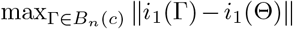is bounded by an integrate function under Condition 2. By the strong law of large numbers, 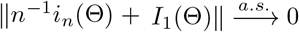These results imply that

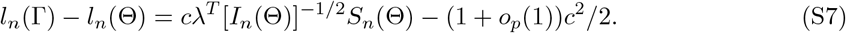

Note that max_*λ*_{*λ*^*T*^ [*I*_*n*_(Θ)]^−1*/*2^*S*_*n*_(Θ)} = ‖*I*_*n*_(Θ)^−1*/*2^*S*_*n*_(Θ) ‖ with

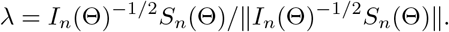

Therefore, (S5) follows from (S7) and

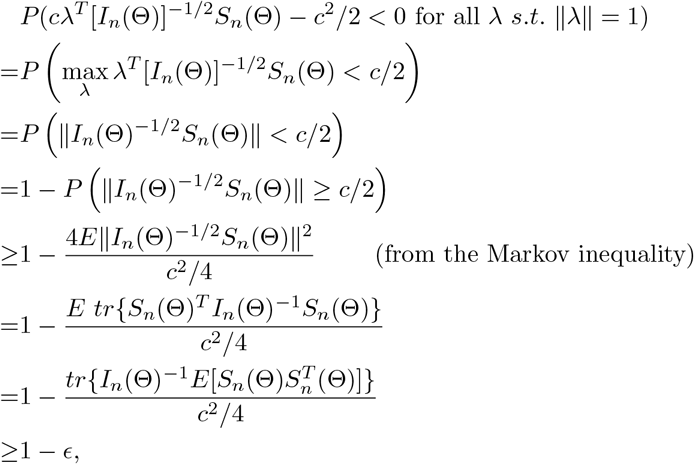

from choosing *c* such that 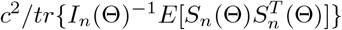 is large enough, which is possible because

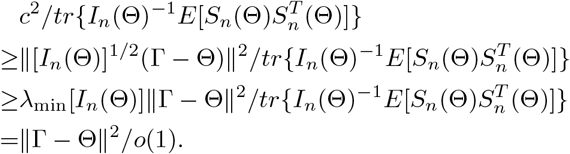

For *n* ≥ *n*_0_, 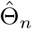 is unique because from Condition 2, the Fisher information *I*_*n*_(Γ) is positive definite in a neighborhood of Θ.

(b) Using the mean value theorem for vector-valued functions, we obtain that

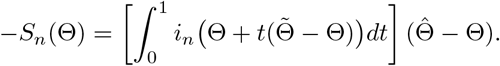

Note that using a similar argument as in (S6), when 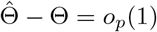,

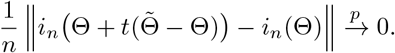

Since 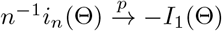 and *I* (Θ) = *nI* (Θ),

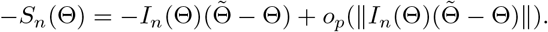

This and Slutsky’s theorem imply that 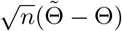 is asymptotically equivalent with

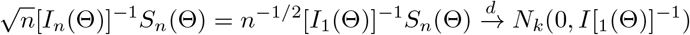

by the central limit theorem for i.i.d. samples. □

### Empirical guidelines for asymptotics

In practice, it is important to know what value of 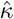 would be considered large enough for the CMLE to perform well, i.e., to have small bias and adequate coverage probability, in presence of many weak invalid IVs.

We follow the same setting as the second simulation study in Section 3 of the main article. We consider two scenarios: (i) *n* = 10^5^, *p* = 20 in Figure 3; and (ii) *n* = 10^6^, *p* = 5 in Figure 4. For each of the 1000 simulation runs, we plot 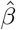 against 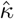. We also plot the two standard error bands centered at *β*_0_ (shaded area). From Figures 3–4, 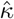 are all larger than 10, and 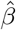 are inside the shaded area, close to the true value *β* = 0.8. Note that because 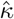 (which relates to the Hessian matrix) is a function of 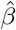, we see that 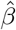 is not symmetric around the true value *β* = 0.8. But this usually does not affect using 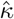 as a condition to evaluate whether the CMLE would perform well. We have also simulated extensively under other settings, we find that the CMLE has good performance when 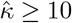.

**Figure 3:**
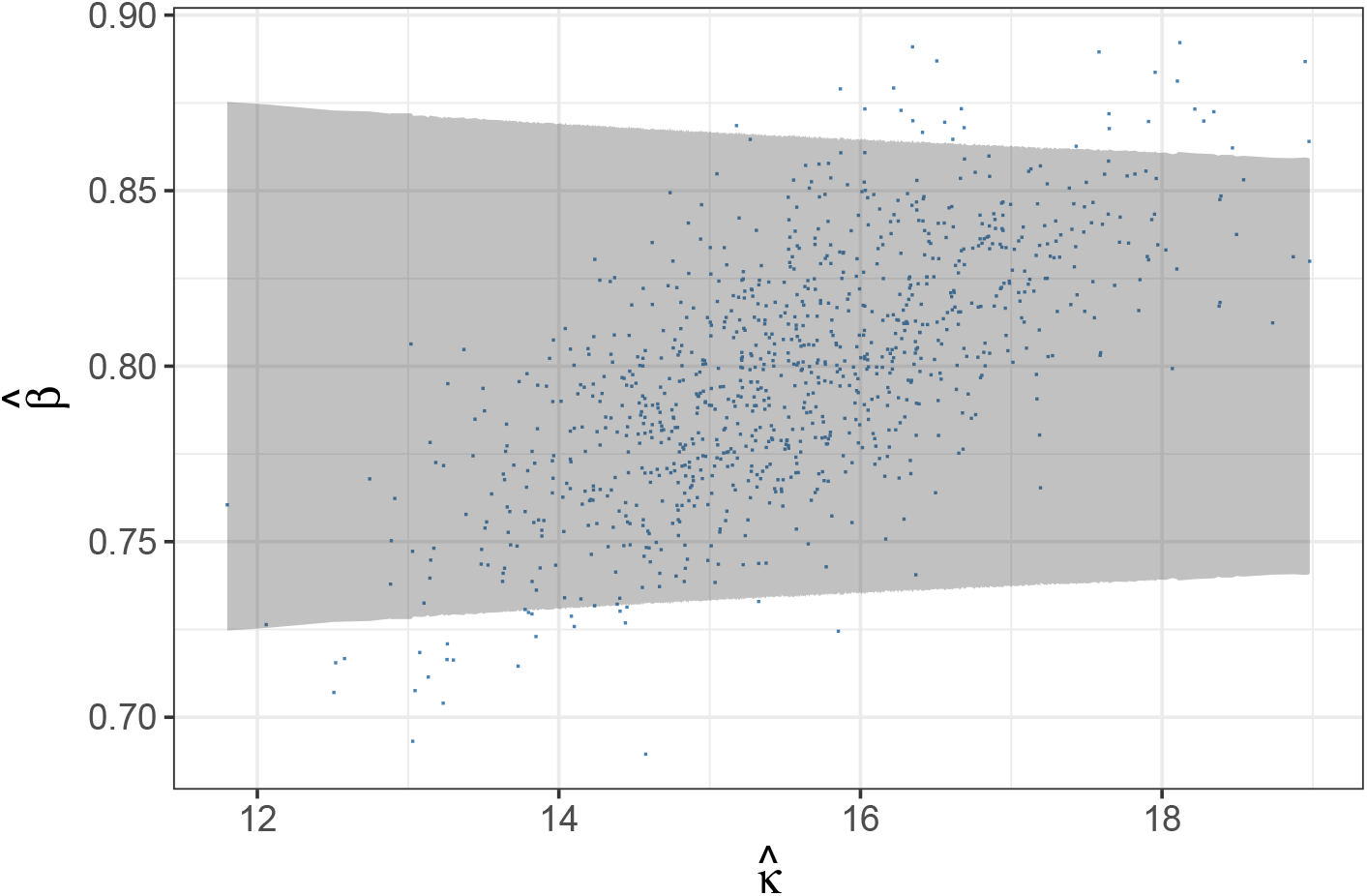
Evaluation of the consistency and asymptotic normality condition for the CMLE under the same setting as the second simulation study in Section 3 of the main article, with *n* = 10^5^, *p* = 20. The x-axis plots the condition 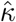, and the y-axis plots values of the CMLE in 1000 simulations. The shaded area represents the two-sided error bands centered at *β* = 0.8.

**Figure 4:**
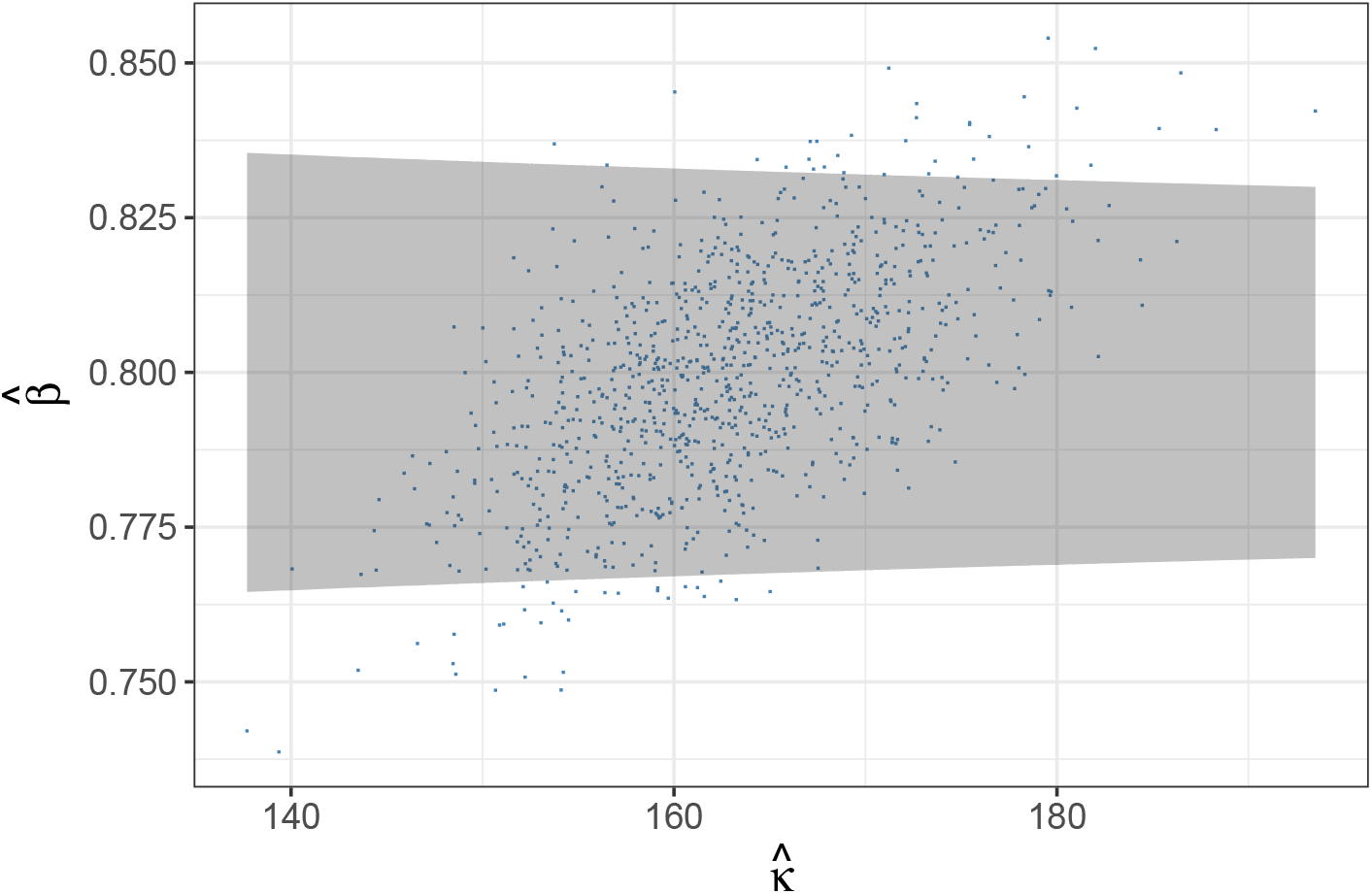
Evaluation of the consistency and asymptotic normality condition for the CMLE under the same setting as the second simulation study in Section 3 of the main article, with *n* = 10^6^, *p* = 5. The x-axis plots the condition 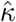, and the y-axis plots values of the CMLE in 1000 simulations. The shaded area represents the two-sided error bands centered at *β* = 0.8.

### Estimation method under Gaussian mixture error

We describe the alternating optimization procedure with user-specified *K* and parametric models *σ*(*Z* = *z*; *η*) and *E*(*Y* | *A* = 0, *Z* = *z*; *θ*) ≡ *µ*_0_(*z*; *θ*) indexed by finite-dimensional parameters *η* and *θ* respectively.

i. Initialization step: maximize the standard Gaussian location scale model

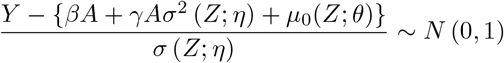

with respect to (*β, θ, η, γ*) by maximum likelihood estimation, to obtain 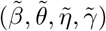. Construct the standardized residuals

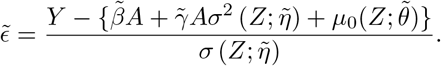
ii. Maximize the Gaussian mixture location scale model

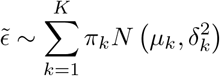

with respect to (*π*_1_, …, *π*_*K*_, *µ*_1_, …, *µ*_*K*_, *δ*_1_, …, *δ*_*K*_) using constrained maximum likelihood estimation to obtain 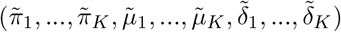. Construct

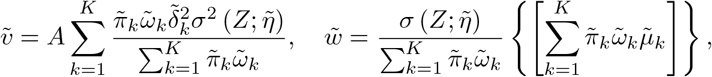

where

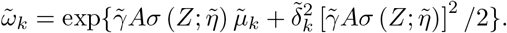
iii. Minimize 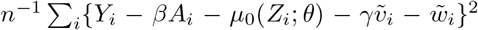 using the least squares method with offset 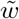 to obtain new 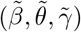, followed by regressing the squared residuals to obtain new 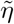. Construct the standardized residuals

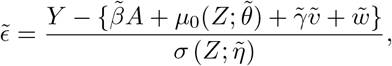

based on the new values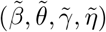
iv. Iterate between steps (ii) and (iii) until change in log-likelihood derived at step (ii) falls below a user-specified tolerance level. In this simulation we iterate until |log *LL*_*j*_ − log *LL*_*j*−1_ |*/*| log *LL*_*j*−1_ *<* 0.001 at the *j*-th iteration.

Asymptotic variance is estimated based on standard M-estimation theory by stacking the estimating functions in steps (ii) and (iii), evaluated at the final parameter values at convergence.

### Semiparametric three-stage estimation

Consider the general location-scale model

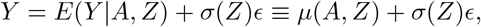

where *ϵ* ⊥ *A, Z*. Under assumptions (B1)-(B3), the conditional mean function is given by

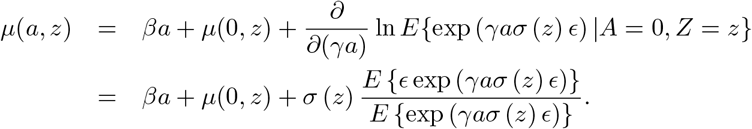

A semiparametric three-stage estimator of (*β, γ*) may be implemented via the following steps:

i. Fit the regression model *µ*(*a, z*) = *m*_*a*_ (*a*)+*m*_*z*_ (*z*)+*m*_*az*_ (*a, z*), where 0 = *m*_*z*_ (0) = *m*_*az*_ (*a*, 0) = *m*_*az*_ (0, *z*) for identification, using a nonparametric method such as generalized additive model. For example, if the support of *Z* is 0, 1, 2, then the nonparametric model for the outcome is of the form 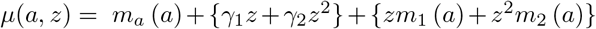 with *m*_1_ (0) = *m*_2_ (0) = 0; a saturated model may be considered if *A* also has finite support. Let 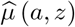 denote the resulting estimator of the mean *µ*(*a, z*).
ii. Using the residuals from (i), fit a nonparametric model for the conditional variance *σ*^2^ (*z*). If the support of *Z* is {0, 1, 2}, then we can specify the saturated model 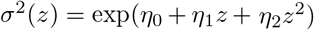. Let 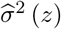 denote the resulting estimator of *σ*^2^(*z*).
iii. Define 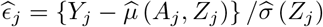 and let

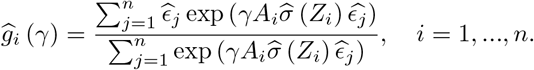 The proposed semiparametric three-stage estimator of (*β, γ*) is

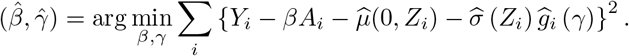

We note that a potentially more efficient approach is to maximize the log-likelihood for the model using a kernel estimator for the density of standardized residuals from step (iii). Specifically, let

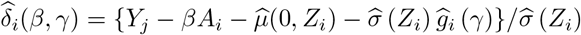

and define

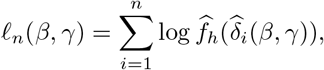

where 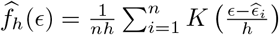is a kernel density estimator with bandwidth *h >* 0. The alternative estimator of (*β, γ*) is given by

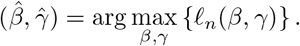

